# Genome-wide discovery reveals 30 loci for choroidal thickness and uncovers potential causal links with angle-closure glaucoma

**DOI:** 10.64898/2026.05.26.26354075

**Authors:** Samantha Sze-Yee Lee, Carol A Wang, Victor A de Vries, Dirk J van Hemert, Alicia Schulze, Caroline Brandl, Asma M Aman, David Alonso-Caneiro, Hélène Choquet, Mathias Gorski, Christopher J Hammond, Iris M Heid, Michael L Hunter, Pirro Hysi, Chen Jiang, Jost B Jonas, Caroline CW Klaver, Sander CM Kneepkens, Simon C König, Gareth Lingham, Christian Luber, Philip E Melton, Craig E Pennell, Wishal D Ramdas, Scott A Read, Alexander K Schuster, Ya Xing Wang, Martina E. Zimmermann, International Glaucoma Genetics Consortium, Anthony P Khawaja, Puya Gharahkhani, Stuart MacGregor, Jeremy A Guggenheim, David A Mackey

**Affiliations:** The University of Western Australia, Centre for Ophthalmology and Visual Science (incorporating the Lions Eye Institute), Perth, WA, Australia; School of Optometry and Vision Science, UNSW, Sydney, New South Wales, Australia; QIMR Berghofer, Brisbane, QLD, Australia; School of Medicine and Public Health, The University of Newcastle, Newcastle, NSW, Australia; Reproductive and Family Health Research Program, Hunter Medical Research Institute, New Lambton Heights, NSW, Australia; Department of Ophthalmology, Erasmus University Medical Center, Rotterdam, The Netherlands; Department of Epidemiology, Erasmus University Medical Center, Rotterdam, The Netherlands; The Generation R Study Group, Erasmus University Medical Center, Rotterdam, The Netherlands; Institute of Medical Biostatistics, Epidemiology and Informatics, University Medical Center of the Johannes Gutenberg-University of Mainz, Mainz, Germany; Department of Ophthalmology, University Hospital Regensburg, Regensburg, Germany; Department of Genetic Epidemiology, University of Regensburg, Regensburg, Germany; Faculty of Health, Medicine and Behavioural Sciences, The University of Queensland, Brisbane, QLD, Australia; School of Science, Technology and Engineering, University of Sunshine Coast, Petrie, QLD, Australia; Centre for Vision and Eye Research, Optometry and Vision Science, Queensland University of Technology, Brisbane, QLD, Australia; Kaiser Permanente, Division of Research, Pleasanton, CA, USA; Department of Health Systems Science, Kaiser Permanente Bernard J. Tyson School of Medicine, Pasadena, CA, USA; Departments of Ophthalmology and Twin Research & Ageing, Genomic Epidemiology, King’s College London, London, United Kingdom; School of Population and Global Health, Faculty of Health and Medical Sciences, University of Western Australia; Busselton Population Medical Research Institute, Busselton, Western Australia, Australia; Rothschild Foundation Hospital, Paris, France; Singapore Eye Research Institute, Singapore National Eye Center, Singapore; Beijing Visual Science and Translational Eye Research Institute (BERI), Beijing Tsinghua Changgung Hospital, Tsinghua Medicine, Tsinghua University, Beijing, China; LV Prasad Eye Institute, Hyderabad, Telangana, India; Department of Ophthalmology, Radboudumc, Nijmegen, The Netherlands; Institute of Molecular and Clinical Ophthalmology Basel, Basel, Switzerland; Department of Ophthalmology, University Medical Center of the Johannes Gutenberg-University Mainz, Mainz, Germany; Centre for Eye Research Ireland, Sustainability and Health Research Hub, Technological University Dublin, Dublin, Ireland; Centre for Eye Research Australia, Melbourne, Australia; Menzies Institute for Medical Research, School of Medicine, University of Tasmania, Hobart, TAS, Australia; NIHR Biomedical Research Centre, Moorfields Eye Hospital NHS Foundation Trust & UCL Institute of Ophthalmology, London, UK; School of Medicine, University of Queensland, Brisbane QLD, Australia; School of Optometry & Vision Sciences, Cardiff University, Cardiff, United Kingdom

**Keywords:** choroid, genome-wide association studies, macular degeneration, Mendelian randomisation, primary angle-closure glaucoma, refractive error, Canadian Longitudinal Study on Aging

## Abstract

The choroid is critical for maintaining vision and implicated in several ocular diseases, being the sole source of nutrients and waste removal for the outer retina. Genetic discovery can help elucidate the pathways through which choroidal features influence disease risk. Our meta-analysis of genome-wide association studies (n= 78,682 participants) identified 30 genomic regions, including 20 novel loci, associated with choroidal thickness. Findings suggest inflammatory and vascular processes drive choroidal thickness, with overlapping mechanisms shared with refractive error. Genome-wide independently significant SNPs accounted for 18.7% of the genetic variance in choroidal thickness. Mendelian randomisation analyses showed a causal effect of age-related macular degeneration on choroidal thickness, and suggest a bidirectional causal effect between choroidal thickness and primary angle-closure glaucoma. These findings provide insight into the shared genetic architecture and biological pathways linking choroidal thickness and related diseases.

---

The choroid is a vascular-rich layer sandwiched between the retina and sclera; in mammals, it serves as the only source of nutrient and waste removal for the highly metabolic outer retina, including the photoreceptors. Because of its vascular nature, choroidal thickness (ChT) varies with chronic systemic or ocular disease, especially those that involve vascular or inflammatory processes. The choroid tends to be thinner in age-related macular degeneration (AMD)^1^ and pathological myopia;^2^ but thicker in polypoidal choroidal vasculopathy^1^ and primary angle-closure disease.^3,4^ Associations between ChT and best-corrected visual acuity have also been found in population-based cohorts,^5,6^ prompting the suggestion that ChT could be a biomarker of visual function.

Despite the importance of the choroid, its genetics has received little research attention. Studies in Asian populations have noted associations between ChT and variants within *CFH*, a major AMD susceptibility gene.^7–9^ A study of 38,000 Europeans in the UK Biobank, revealed 19 lead SNPs across 16 genomic risk loci for ChT and a SNP-based heritability of 16.8%. Together, these studies suggest that the ChT has a heritable, polygenic component. None of the previously reported loci have been independently replicated. In the current study, we report 20 novel genetic loci for ChT. We further explored causal relationships of ChT and genetic architecture overlaps with refractive error, AMD, and primary angle-closure glaucoma (PACG).

## Results

### Stage 1 (discovery) GWAS and replication

In Stage 1, we meta-analysed genome-wide association studies (GWAS) of ChT comprising 33,859 participants of European descent from the International Glaucoma Genetics Consortium (IGGC), Generation R (GenR),^10^ and the Canadian Longitudinal Study on Aging (CLSA).^11,12^ Cohort details are given in **Supplementary Table 1**. In the IGGC and GenR cohorts, ChT was directly phenotyped using optical coherence tomography (OCT) scans followed by manual or automated segmentation and thickness measurement. In the CLSA, ChT was estimated from fundus photographs by a machine-learning algorithm (see Methods). Raw ChT measures underwent a rank-inverse normal transformation prior to analysis. For the analyses of all cohorts except CLSA, the ChT GWAS analyses were adjusted for age, sex, the first 10 principal components, and either axial length or spherical equivalent refractive error. For the CLSA, the GWAS analysis was initially adjusted for age, sex, and the first 10 principal components; subsequently, spherical equivalent refractive error was adjusted for using mt-COJO (see methods). Meta-analysis was performed using an inverse variance fixed-effect scheme.

As shown in **Figure 1** and **Supplementary Table 2**, 6 discrete genomic regions were identified at genome-wide significance in the Stage 1 analysis, including 2 novel loci. All but one (lead SNP rs1651269 in chromosome 5) of the 6 loci were previously linked to refractive error.^13^ The most strongly associated SNP was rs1129038, a variant in the 3’ untranslated region of *HERC2*. Median I^2^ was 0, suggesting minimal heterogeneity.

**Figure 1.**
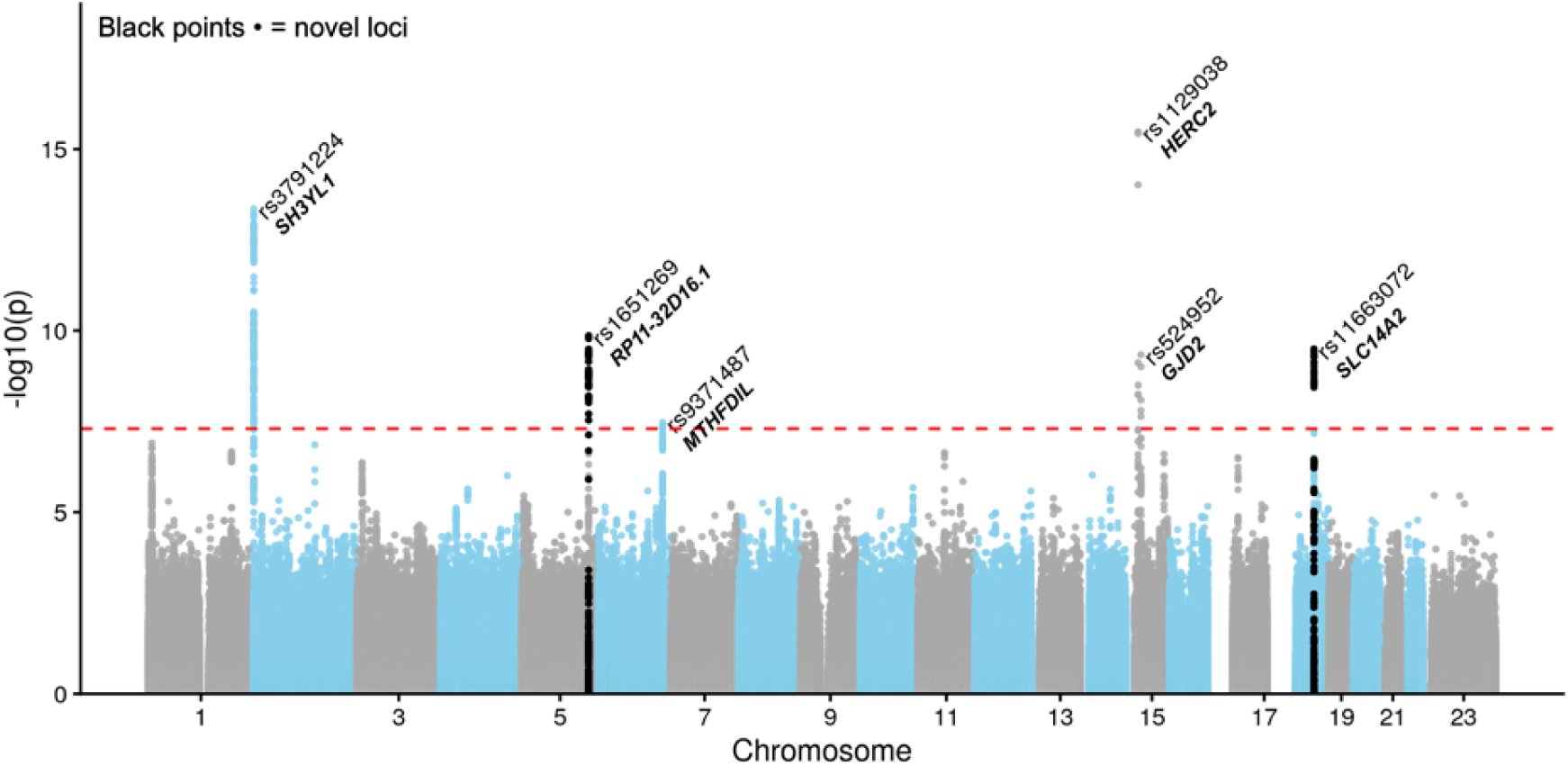
Manhattan plot of the Stage 1 GWAS meta-analysis (n=33,516 participants). Lead SNPs are annotated along with the closest gene. Black points indicate novel loci. Genome-wide significance line (red) at P<5e^-08^.

Of the 19 lead SNPs previously reported in the UK Biobank GWAS of ChT,^14^ we replicated 10 at a Bonferroni-corrected significance level of *P*< 0.0026 (0.05/19) (**Table 1**), while 3 loci were nominally replicated at p< 0.05. All but one locus (lead SNP rs707739, p= 0.90 in the Stage 1 GWAS) had the same direction of effect between the UK Biobank and the Stage 1 GWAS results. The two SNPs associated with ChT in Asian ancestry samples reported by Hosoda et al.,^8^ were not associated with ChT in our Stage 1 GWAS, nor were proxy variants within 500kb, despite a relatively high frequency of the minor allele in both SNPs (20–33%).

### Stage 2 (discovery + UK Biobank) GWAS meta-analysis

We next meta-analysed the Stage 1 GWAS with the UK Biobank^14^ GWAS summary statistics (total n= 78,682). Conditional analyses using FUMA identified 91 significantly independent SNPs (**Supplementary Table 3**) including 35 lead SNPs across 30 genomic regions, 20 of which were novel (**Figure 2** and **Table 2**). Heterogeneity was minimal (median I^2^=0).

**Figure 2.**
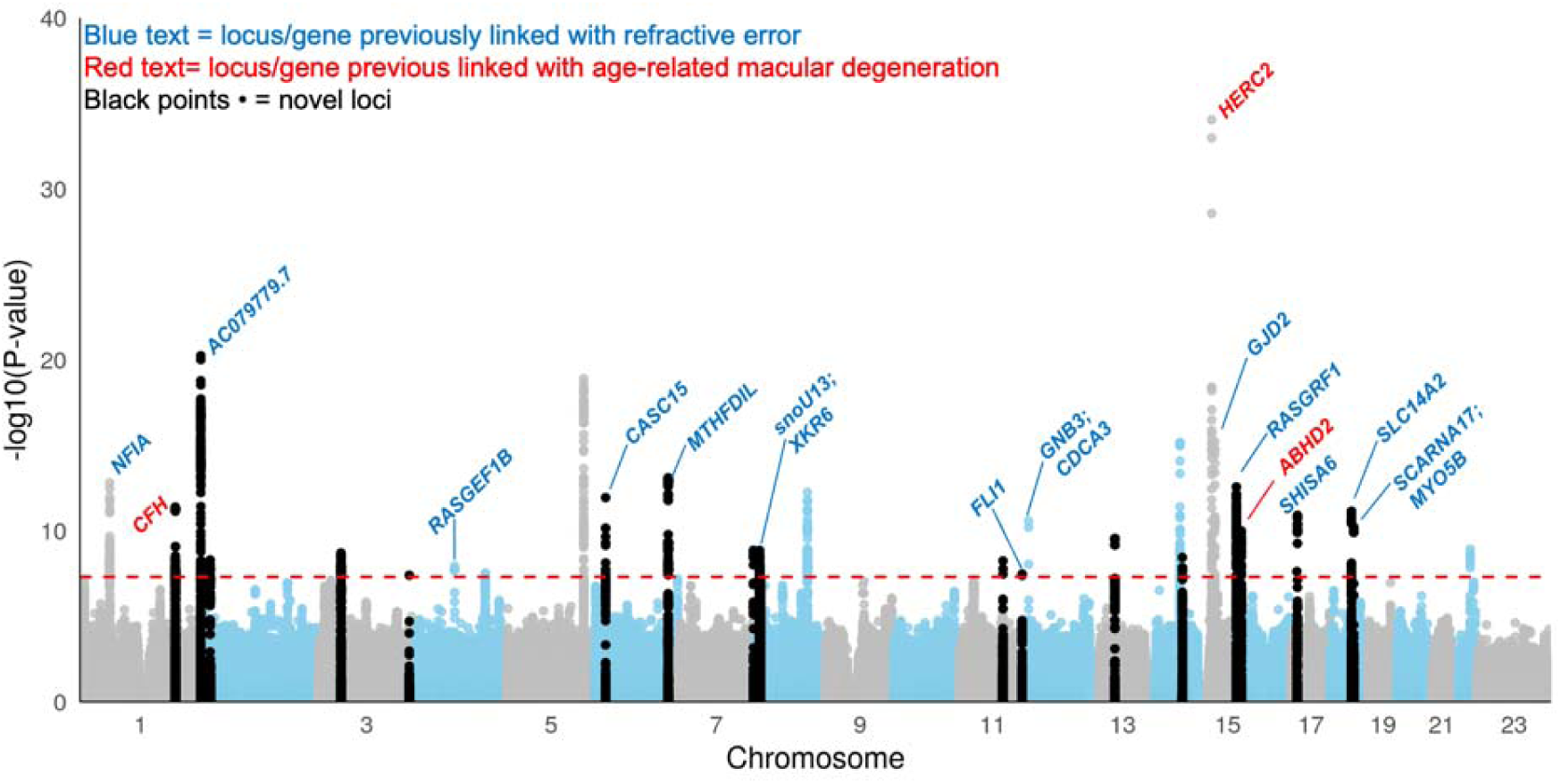
Manhattan plot of Stage 2 (combined) GWAS meta-analysis comprising 72,344 participants. The nearest gene at each locus is annotated. Loci previously linked with refractive error or its endophenotypes (blue) or age-related macular degeneration (red) are indicated. Black points indicate novel loci. Genome-wide significance line (red) at P<5e^-08^.

Given the variation in phenotyping method between the IGGC+GenR (OCT imaging with full access to the chorioscleral interface), CLSA (machine-learning estimated), and UK Biobank (OCT imaging with limited depth), we calculated the genetic correlation for the ChT between these cohorts using linkage disequilibrium score regression (LDSC). Genetic correlation of IGGC+GenR with either CLSA or UK Biobank were high (∼0.99; *P* ≤ 0.001), and moderate between the CLSA and UK Biobank (r_g_= 0.76 [95%CI= 0.10], *P*= 7.4×10^-11^). The betas of the SNPs that reached genome-wide or suggestive significance (*P*< 1×10^-5^) in the combined GWAS were strongly correlated between all three cohorts (**Supplementary Figure 1**).

### Top variant for ChT, rs12913832

Variants within *HERC2* remained the most strongly associated with ChT in the Stage 2 meta-analysis, with an intronic lead SNP rs12913832 (*P*= 9.08×10^-35^). *HERC2* encodes a ubiquitin ligase that is expressed widely in the body and plays a critical role in DNA repair by marking other proteins for degradation. This gene has wide ranging effects given its large size, and has been linked with AMD,^15,16^ intraocular pressure,^17^ as well as eye, skin, and hair pigmentation.^18,19^ The latter traits are influenced by the lead SNP, rs12913832, via regulating expression of *OCA2*, a well-known gene for melanin synthesis.^18,19^ Furthermore, each additional copy of the A allele in rs12913832 is linked to darker irides and confers a ∼1.3 increased odds of PACG.^15,20^ Contrary to reports of thicker choroids in eyes with primary angle-closure, we observed that the A allele in rs12913832 is associated with a thinner choroid (**Table 2**).

### Genetic overlap with AMD and other macular traits

In addition to variants within *HERC2*, an intronic SNP, rs1329427, near *CFH* was identified; this SNP is independent of rs3753394, located 2kb upstream of *CFH,* reported by Hosada et al.^8^ Another implicated AMD-associated locus was found near *ABHD2*, a gene that modulates lipid metabolism^21^ and may interact with mitochondrial DNA variants to influence AMD risk.^22^ Notably, a single exonic variant, rs5442 (*P*= 2.64×10^-11^) was identified which exhibited a high CADD (combined annotation dependent depletion) score of 26.5. Located within *GNB3* and *CDCA3*, rs5442 also carries signals for retinal vasculature measures,^23^ macular thickness,^24^ and refractive error.^13,25^

### Genetic overlap with refractive error

In addition to rs5442, a further 12 of the 30 lead ChT loci have been implicated in refractive error development (**Table 2**). Of these 13 regions (including rs5442), the intergenic locus near *GJD2* has the strongest association with myopia.^25^ *GJD2* encodes a gap junction protein that is highly expressed in the retina and facilitates neuronal signalling between retinal cells.^26^ Another potentially deleterious variant, rs2908972 near *SHISA6* (CADD score= 15.7), is enriched in the brain and interacts with AMPA receptors to facilitate neuronal communication.

Variants were also found near or within *XKR6*, which may have a role in apoptosis.^27^ *XKR6* is implicated in several cardiometabolic^28^ and autoimmune diseases such as systemic lupus erythromatosus.^28,29^ The lead variant, rs11250098, is in LD with rs656319 which has been reported to have suggestive evidence of association with pathological myopia.^30^

### Genetic overlap with other phenotypes

Another key variant is the intergenic lead SNP rs135173 (CADD score= 14.5), located near *PDGFB*, which encodes a protein that promotes capillary tube formation by signaling the proliferation and recruitment of mural cells to the embryonic vasculature and maintaining pericyte coverage, thus playing a crucial role in angiogenesis and blood vessel stabilisation.

Variants were additionally found near or within genes associated with glaucoma endophenotypes (e.g., *MTHFD1L* and *AC092594.1* genes for optic disc features;^31^ *TYR* for intraocular pressure),^17^ corneal biometrics (e.g., *CASC15*, *HERC2*),^32^ and corneal disease (*CFH* for Fuch’s corneal dystrophy).^33^ Other common phenotypes that have been linked with several of the identified loci include measures of blood lipid levels,^34^ blood or pulse pressure,^35^ and risk of type 2 diabetes (**Table 2**).

### Functional mapping

Using expression quantitative trait locus (eQTL) mapping, we explored how the Stage 2 genome-wide significant variants may impact gene expression in 48 tissues within the GTEx v8 database and the retina (EyeGEx). A total of 104 genes had significant eQTL effects across the 49 tissues (**Supplementary Figure 2**). **Supplementary Figure 3** shows a heatmap of the effect size of significant eQTLs genes, at false discovery rate [FDR] < 0.05) in 16 tissues with selected properties relevant to the choroid (light-responsiveness, vascular control, or immune responses). eQTL regulating expression of *SH3YL1* were significant across most tissues, including the retina, although it should be noted that this gene is expressed throughout the body. The most significant eQTL effects in the retina related to the expression of *RASGRF1*, which is a genomically imprinted gene associated with refractive error.^36^

Chromatin interaction mapping suggested that 83 of the 91 independently significant SNPs physically interacted with target gene promoters across 26 tissues or cell lines (**Supplementary Figure 3**). Notably, the exonic rs5442 variant was associated with significant chromatin interactions at the *GNB3* promoter in the prefrontal cortex,^37^ suggesting a potential role in regulating *GNB3* by altering chromatin structure.

### Gene-based analyses

The Stage 2 GWAS summary statistics were mapped to 20,175 genes. At a Bonferroni-corrected threshold of *P*< 2.48×10^-6^ (0.05/20,175), 29 genes reached statistical significance for association with ChT (**Table 3**). Eighteen of these 29 genes were not annotated to any genome-wide significant SNP in the Stage 2 meta-analysis, including 2 genes coding for CFH-related proteins (*CFHR1* and *CFHR4*). The most strongly associated gene was *SMOC1* (*P*= 1.42 ×10^-15^), which encodes a basement membrane-enriched proteins expressed during embryonic development, facilitating cell differentiation and growth. Other top genes were *SH3YL1* (*P*= 1.41×10^-13^), *CFH* (*P*= 2.26×10^-13^), *MTHFD1L* (*P*= 2.25×10^-11^), and *MSRA* (*P*= 41.21×10^-10^). The latter, which encodes the methionine sulfoxide reductase-A enzyme, has been previously linked with pathological myopia.^30^

### Pathway analysis

A total of 17,012 pathways, curated from various sources, were tested, but none survived multiple testing correction (*P*< 2.94×10^-6^; 0.05/17,012). At FDR <0.05, 5 pathways – cell-substrate junction organisation, oxidative damage response, chromatin organisation, renal system development, and Alzheimer’s disease pathways – were associated with ChT (**Supplementary Table 4**).

### Polygenic score testing

A polygenic score (PGS) was generated from the genome-wide significant SNPs from the Stage 2 meta-analysis using the clumping and thresholding method,^38^ and evaluated on 609 European participants from Australia, independent of the cohorts used in the Stage 1 and 2 GWAS (**Supplementary Table 1**). Using linear regression analysis corrected for age, sex, genotyping batch, axial length, and the first 5 principal components, each standard deviation (SD) higher in PGS was as expected associated with thicker choroids by 12.6µm (95%CI= 6.3 to 18.9, *P*= 9.7×10^-5^). The PGS explained 1.9% (95%CI= 0.3 to 4.3) of the phenotypic variance. Given the strong age effect on ChT, we repeated these analyses in only participants aged <40 years (n= 512) and showed similar results (ß= +12.13µm per SD increase in PGS; r^2^= 2%). Because of the strong link between ChT and refractive error, we further tested for a cross-trait PGS association with axial length. A weak, non-significant association was observed (ß= -0.07mm per SD increase in PGS; 95%CI= -0.15 to 0.01, *P*= 0.09; **Figure 3**).

**Figure 3.**
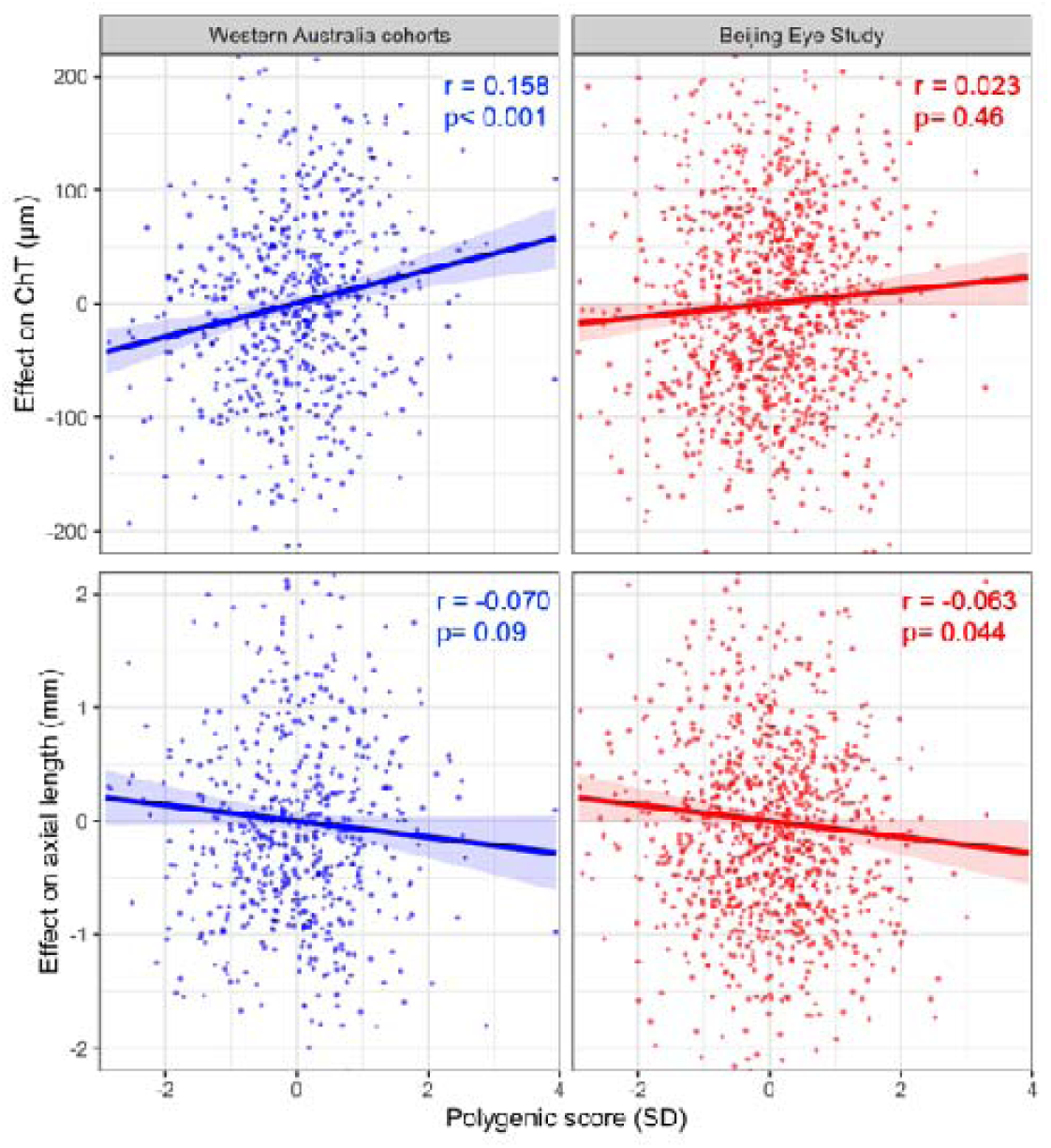
Association between a polygenic score for ChT with either ChT (top row) or axial length (bottom row). The test cohorts comprise participants of European (left panel; blue) or East Asian (right panel; red) genetic ancestry. ChT= choroidal thickness; SD= standard deviation.

We further tested the PGS in 1,128 East Asian participants, aged 50+ years, from the Beijing Eye Study to explore whether the genetic architecture for ChT might be shared across ancestry. The PGS was not significantly associated with ChT in the Beijing Eye Study participants (ß= +2.0µm per SD increase in PGS, *P* = 0.45), although the direction of association was consistent with that in Europeans. However, each SD increase in the PGS was associated with a -0.07mm decrease in axial length in the East Asian sample (95%CI= - 0.14 to -0.00, *P*= 0.044; **Figure 3**).

### Heritability and genetic correlation

The SNP heritability for ChT was 0.13 (95%CI= 0.11 to 0.16) estimated with LDSC. The 35 lead SNPs discovered in the Stage 2 GWAS collectively accounted for 18.7% of the genetic variance for this phenotype.

Using summary statistics from the Stage 2 GWAS meta-analysis and those for refractive error reported by Hysi et al.,^13^ the genetic correlation between ChT and refractive error was estimated at 0.113 (95%CI= 0.043 to 0.183), indicating genetic predisposition to a thinner choroid is linked to more negative refractive error (myopia). We further conducted a genomic relation matrix bivariate linear mixed-effect model with genetic restricted maximum likelihood (GREML) in 2,992 European participants from the Busselton Healthy Aging Study (BHAS). Narrow-sense heritability of ChT was estimated to be 0.38 (95%CI= 0.20 to 0.55). However, the genetic correlation of ChT with either refractive error (Estimate= 0.20 [95%CI= -0.14 to 0.54], *P*= 0.25) or axial length (Estimate= -0.08 [95%CI= -0.37 to 0.21], *P*= 0.57) was not significant in the GREML models. A pedigree-based analysis conducted on 673 individuals from 302 families from various epidemiological cohorts in Western Australia yielded a similar heritability estimate of 0.38 (95%CI= 0.34 to 0.41) and a non-significant genetic correlation with refractive error or axial length (*P*= 0.21 and 0.41, respectively).

GWAS summary statistics for AMD and PACG were obtained from He et al.’s^39^ and Verma et al,^15^ respectively. LDSC-estimated genetic correlation for ChT with AMD and PACG were 0.04 (*P*= 0.37) and 0.02 (*P*= 0.74), respectively. GREML analyses was not conducted for ChT and AMD or PACG due to the low prevalence of these phenotypes in available samples and consequent low statistical power.

### Colocalisation

Colocalisation analysis with approximate Bayes factor performed for each of the 30 genomic regions harbouring lead ChT variants revealed 10 loci with strong evidence of colocalisation with refractive error (PP4 >0.8) and 2 with moderate evidence (PP4= 0.5–0.8) (**Supplementary Table 5**). All but one of these 12 regions have been reported in previous GWAS of refractive error.

The exonic variant rs5442 displayed evidence of colocalisation with both refractive error (PP4 >0.99) and AMD (PP4= 0.72). Two further variants, rs875391 and rs135173, also had strong evidence of colocalisation with AMD (**Table 2**); both have been detected in previous AMD GWAS.^39^ Variants within other known AMD genes – rs1329427 in *CFH*, rs12913832 within *HERC2* – did not show evidence of colocalisation between ChT and AMD. Six variants had strong evidence of colocalisation with PACG (**Supplementary Table 5**), but only rs12913832 has been reported in previous PACG GWAS.

### Two-sample Mendelian randomisation (MR)

We next explored bidirectional causal associations of ChT with refractive error, AMD, and PACG. Instrumental variables (IVs) are shown in **Supplementary Table 6**. The instrumental variables for ChT, refractive error, AMD, and PACG comprised of 23, 49, 43, and 14 SNPs, respectively, explaining 1.4%, 26.7%, 64.8%, and 10.3% of the phenotypic variance, with a minimum F-statistic of ∼30 for each trait-variant pair (**Supplementary Table 6)**. As shown in **Figure 4**, there was no evidence of a causal effect of ChT on refractive error or AMD risk. There was tentative evidence of a causal effect of refractive error on ChT, using the MR-Egger method (*P*= 0.020), with intercept -0.015 (*P*= 0.06) suggesting weak directional pleiotropy (**Supplementary Table 7**). The other MR methods suggested a causal relationship between refractive error and ChT close to the null.

**Figure 4.**
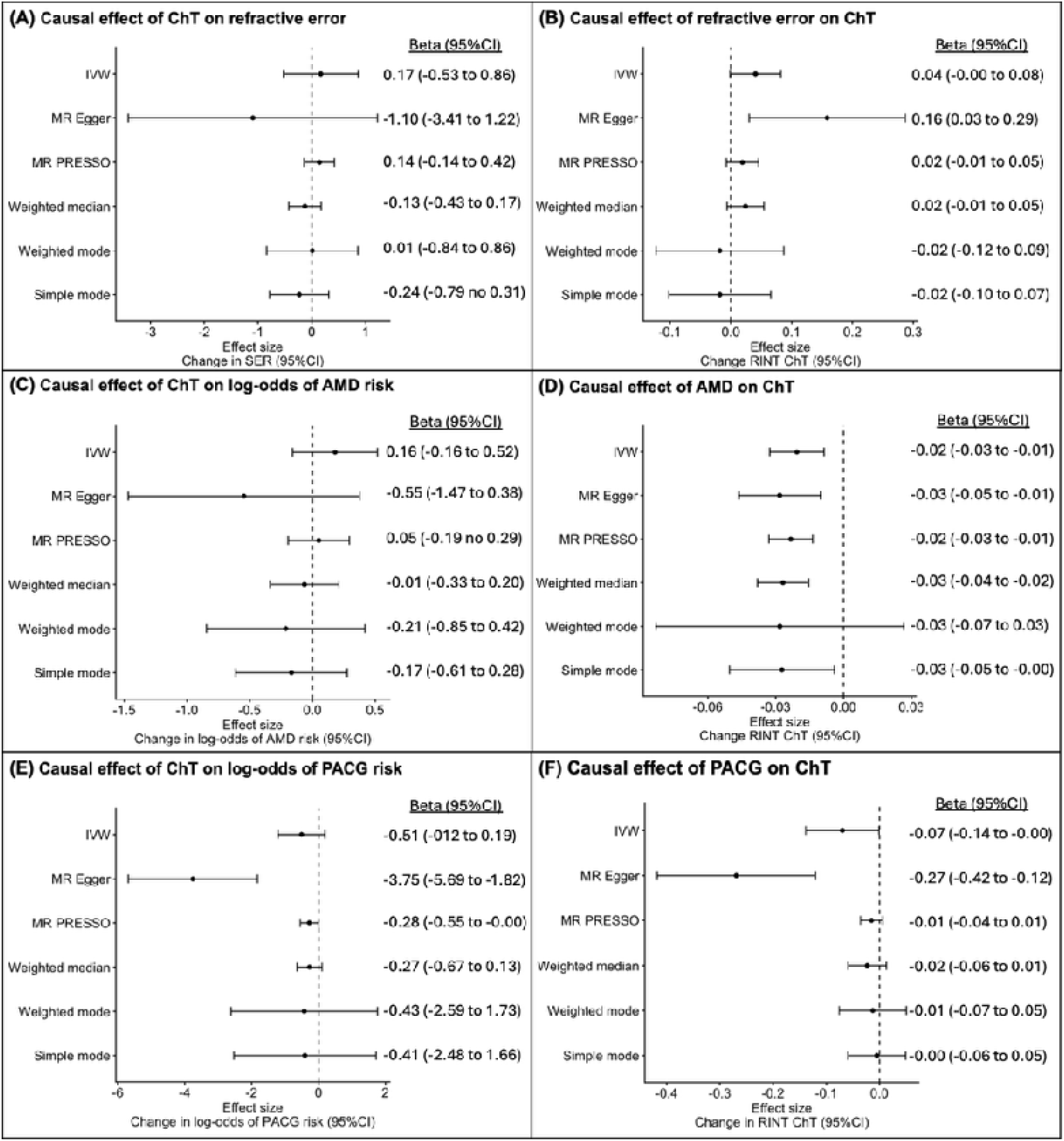
Mendelian randomisation analyses for putative causal effects of (A) ChT on SER; (B) SER on ChT; (C) ChT on AMD risk; (D) AMD on ChT; (E) ChT on PACG risk; and (F) PACG on ChT. AMD= age-related macular degeneration; ChT= choroidal thickness; CI= confidence interval; PACG= primary angle-closure glaucoma; RINT= rank-inverse transformed; SER= spherical equivalent refractive error.

By contrast, there was more consistent evidence of a causal effect of AMD on ChT (**Figure 4**). There was no evidence of pleiotropy, although heterogeneity was high (*P*= 7.58×10^-8^, **Supplementary Table 7**). Leave-one-out sensitivity analysis did not suggest that any specific variant(s) were exerting excessive influence on the MR effect estimates (**Supplementary Figure 4**).

There was some evidence of bidirectional causal effects between PACG and ChT based on the MR-Egger method (**Figure 4**), but this was not significant on most other methods, suggesting pleiotropy may bias the results (**Supplementary Table 7**). Leave-one-out analyses revealed that rs12913832 was the main driver of the observed causal relations on MR-Egger (**Supplementary Figure 5**). We further investigated this using MR-PRESSO, which detected and removed outlying pleiotropic effects in variants rs12913832, rs62186481, and rs973077. Removing these SNPs as instrumental variables showed borderline significance of a causal effect of ChT on PACG risk (*P*= 0.048), while the causal effect of PACG on ChT failed to reach statistical significance (*P*= 0.13).

## Discussion

We identified 20 novel loci associated with ChT, in addition to replicating 10 previously reported genomic regions for this complex trait.^8,14^ Notably, our findings prioritise rs5442 as a high-confidence causal candidate based on its high CADD score, location in the final exon of *GNB3*, and colocalisation with refractive error and AMD. In the eye, *GNB3* has been implicated in refractive error,^13,15,25^ retinal vasculature,^40^ and glaucoma.^41^ This gene encodes a beta subunit of a guanine nucleotide-binding proteins that transmits signals from cell surface receptors to intracellular signaling pathways. Its lead SNP, rs5442, is adjacent to, but in linkage equilibrium with, the intensively-studied synonymous variant rs5443, which has system-wide downstream effects, including inflammatory, vascular, and metabolic changes.^42–44^ These can all conceivably affect ChT but the genomic independence of these 2 SNPs suggests rs5442 could have a distinct but possibly parallel causal pathway influencing the trait.

An intronic locus within *CFH*, a pivotal gene as regards to AMD risk, was also identified in the Stage 2 GWAS meta-analysis. Gene-based analyses further implicated complement factor-related genes in ChT. *CFH* is highly expressed in the choriocapillaris,^45^ and complement system activation and inflammation could potentially lead to choriocapillary leakage and oedema. This corroborates observations of choroidal thickening during active inflammation.^46^ However, repeated or chronic inflammation is associated with a thinner choroid in otherwise healthy individuals^47^ as well as those with autoimmune disorders,^46^ which hints that fibrosis and vascular stress causes loss of choriocapillaris.^46^

The most strongly associated variants were located in *HERC2*; specifically, the A allele in lead SNP, rs12913832, is linked with thinner choroids. This seemed to contradict expectations that iris pigmentation or PACG risk, each of which are increased with the A allele at rs12913832,^15,18–20^ is associated with thicker choroids.^3,4,48,49^ While darker irides tend to have higher volume and thus thicker because of melanin accumulation in the tissue;^50^ this does not necessarily apply to the choroid. In angle-closure, we propose that restriction on conventional outflow leads to greater reliance on the uveoscleral outflow; there is evidence that the uveoscleral outflow is modestly pressure-dependent, though to a lesser extent than the conventional outflow.^51^ We posit that an increase in uveoscleral outflow could be reflected by thicker choroids as aqueous is absorbed by choroidal vessels from the suprachoroidal space. On the other hand, persistently thinned choroids could indicate limited aqueous absorption into the choroidal vessels, leading to glaucoma. Arora et al.^48^ reported that participants with angle-closure, regardless of glaucoma status, had thicker choroids (318µm) than controls or those with open-angle glaucoma (∼234µm). However, those with PACG had thinner choroids (272µm) than participants with primary angle closure suspects and primary angle-closure without glaucoma (319–357µm). Mual et al.^49^ observed that participants with PACG had thinner choroids than those with open-angle glaucoma, while angle-closure without glaucoma had 100µm-thicker choroids than those with PACG or suspected open-angle glaucoma. However, these previous studies had very small samples (a few dozen per group)^4,48,49^ thus no definitive conclusions could be drawn.

Nonetheless, these observations suggest that choroids are thickened in primary angle-closure only when there is no glaucoma. We posit that the A allele in rs12913832 could trigger an initial effect on ChT or PACG risk, creating a positive feedback loop where IOP elevation further restricts choroid’s ability to expand. However, any such effect of ChT on PACG risk would be conditional on the presence of angle closure, which is relatively uncommon in the population; this may explain why MR failed to consistently capture a bidirectional causal relationship. This ChT–glaucoma causal relationship in angle-closure remains speculative and should be investigated in future work.

MR also revealed a causal effect of AMD on choroidal thinning, with largely consistent effects across methods. By contrast, there was little evidence of a causal effect of ChT on AMD risk. There were also specific SNPs that showed strong evidence of colocalisation for AMD and ChT; however, there was minimal genetic correlation for variants across the genome, suggesting that AMD and ChT are genetically distinct for the most part. The link between AMD and ChT is further supported by the gene-set analysis, which provided evidence that vascular and stress response processes affect ChT. The highly vascular nature of the choroid makes it susceptible to hemodynamic and mechanical strain. Our findings suggest a role for cell-substrate junction organisation, specifically cell–extracellular matrix interactions and adhesion dynamics in vascular tissues. These could affect cell adhesion and signalling which are important for tissue integrity and vascular permeability. Oxidative damage response is also a potentially key pathway for modulating ChT.

Approximately half of the loci associated with ChT were shared or showed evidence of colocalisation with refractive error, suggested shared biological mechanisms between these traits. These traits had a modest genetic correlation, although this was not statistically significant across all methods, potentially due to limited statistical power. These observations align with the consistently strong phenotypic associations between thinner choroids and more negative (myopic) refractive error.^52,53^ It has been assumed that eye elongation in myopia stretches and thins the choroid.^54^ Some studies have observed that choroid thinning may precede myopia onset^55^ or progression,^5^ after controlling for baseline refractive error.

However, such conclusions may be influenced by the definition typically used to define myopia onset (spherical equivalent refraction ≤-0.5 D), whereas axial elongation in eyes developing myopia is now known to begin at refractive errors around +1.00 D.^56^ The shared genetic architecture of ChT and refractive error may have led to confounded relationships which our MR analyses struggled to disentangle. Weak evidence of a causal effect of refractive error on ChT was observed in the MR-Egger analysis, which was not supported by the other MR methods. Based on the low variance explained by the IVs for ChT and the mixed results for the causal effect on refractive error on ChT, we interpret these results as suggestive, rather than definitive; this is an important direction of research for future studies.

In independent cohorts, the PGS was predictive of ChT in participants of European ancestry but not those of East Asian ancestry. A lack of cross-ancestry portability is a common feature of PGSs and could have arisen as a result of ancestry-specific patterns of LD rather than a difference in ChT biology.^57^ Somewhat paradoxically, our PGS for ChT was weakly predictive of axial length in the East Asian cohort, with a similar effect size to that seen in the European cohort (the latter result failed to reach statistical significance, potentially due to reduced statistical powe). Lack of statistical power also prevented us from definitively assessing the genetic correlation between ChT and axial length across ancestry groups.

A key strength of the current study was the large sample size (n= 78,682) of the Stage 2 meta-analysis achieved by combining various cohorts including the UK biobank. By doing so, we more than doubled the number of genetic loci known to be associated with ChT. Nonetheless, the current study had a few limitations which should be addressed in future work. First, only European participants were included in the GWAS meta-analysis. While this minimised population stratification effects, this limited the generalisability of the findings and may have contributed to the poor accuracy of the PGS in other ancestry. Second, ChT was measured directly from OCT scans in the IGGC and GenR cohorts, while in the CLSA, ChT was estimated from fundus images via a machine-learning algorithm. Even though OCT imaging was conducted in the UK Biobank cohort, the imaging device used was nat optimised for choroidal imaging and hence unable to traverse thicker choroids, resulting in a ceiling effect where higher ChTs were underestimated.^14^ Nonetheless, the genetic correlation of ChT between cohorts was high, suggesting that phenotyping heterogeneity did not have a major adverse impact. Most SNPs associated with ChT in the original UK Biobank GWAS were replicated in our Stage 1 GWAS, further confirming the validity of ChT measures across cohorts. Third, only the subfoveal ChT was studied. Imaging the peripheral choroid and quantifying ChT across the globe using OCT remains challenging; we thus restricted our analysis to just the central choroid. Finally, genetic variants associated with ChT potentially have differential effects depending on age. Because the current GWAS cohorts comprised mostly older adults, we may have missed genetic variants associated with ChT at a younger age. Nonetheless, our PGS for ChT was validated in an independent cohort of young adults.

This study builds on prior genetic studies of ChT by doubling the number of loci associated with the trait. Yet, the lead variants identified accounted for less than one-fifth of the genetic variance in ChT, suggesting that future studies with larger samples or that assess rare variants will reveal further loci. With advancements in imaging technology and analysis methodologies, access to ChT measurement in large clinical or population-based cohorts will be a promising direction for future research. This will enable greater statistical power for genetic discover, allowing robust Mendelian randomisation analyses on ChT’s causal relations with refractive error, PACG, and other chorioretinal diseases.

## Methods

### Study design

This study comprises 2 stages of GWAS, post-GWAS analyses on the Stage 2 summary statistics, PGS testing, and MR. The Stage 1 GWAS meta-analysis included 33,516 European participants from the (IGGC), Generation R (GenR), and the Canadian Longitudinal Study of Aging (CLSA) cohorts. The IGGC cohorts included in this study are the AugUR Study, BHAS, Rotterdam Study, Gutenberg Health Study, and the Raine Study Gen2.

The AugUR Study^58^ is a cohort study comprising elderly individuals age 70+ years in the city and county of Regensburg, Germany. Eligible residents were identified by local registries and invited to participate in the baseline 3-hour study program at the Regensburg University Hospital, including OCT imaging. All participants provided written informed consent. The study protocol was approved by the University of Regensburg Ethics Committee and complied with the Declaration of Helsinki.

The BHAS^59^ comprised middle-aged and older individuals that resided in Busselton, a coastal city in Western Australia. All non-institutionalised adults born 1946-1964 living in the local government based on the electoral roll were invited to participate in the BHAS. At Phase 1 of the BHAS (2010–2016), participants underwent autorefraction (ARK-30, Nidek, Japan), while Phase 2 of the study (2016–2022) included OCT imaging, from which the ChT was phenotyped. All health and medical examinations conducted in the BHAS was approved by the University of Western Australia Human Research Ethics Committee (UWA HREC) and complied with the Declaration of Helsinki. All participants provided written informed consent following a full explanation of the nature of the study prior to participation.

The Rotterdam Study^60^ is an ongoing cohort study that started in 1990 in the City of Rotterdam, Netherlands. Participants age 55+ years living in the Ommoord district were recruited in 1990–1993 (RS-I), 2000–2001 (RS-II), 2006–2008 (RS-III), and 2016–2020 (RS-IV). Each wave of recruitment enrolled new participants, in particularly those who had reached the age criterion or moved into the study district. Eligibility of the study were extended to those age 45+ years for RS-III and 40+ years for RS-IV. The Rotterdam Study was approved by the Medical Ethics Committee of the Erasmus MR and the Dutch Ministry of Health, Welfare, and Sport. All participants provided written informed consent and the study was conducted in compliance with the Declaration of Helsinki.

The Gutenberg Health Study^61^ is a population-based cohort study comprising 15,010 individuals randomly drawn from local government registry offices and invited to participate. The baseline examination was conducted in 2007–2012, while the follow-up was conducted in 2012–2017 and included OCT imaging. The study was approved by the Medical Ethics Commission of Rhineland-Palatinate, as well as local and Gutenberg-University of Mainz data protection officials. All participants provided written informed consent prior to participation and the study adhered to the tenets of the Declaration of Helsinki.

The Raine Study^62^ followed a cohort since their births it 1989–1992 at the King Edward’s Memorial Hospital in Perth, Western Australia. This original cohort eventually formed the Gen2 cohort of the study, as their parents (Gen1), grandparents (Gen0), and offspring (Gen3) were eventually recruited into the Raine Study. The Gen participants underwent a series of medical examinations since, including eye examinations at the Gen2 20- and 28-year follow-ups. Ocular biometry (IOLMaster V5, Carl Zeiss Meditec AG) and OCT imaging were done at both follow-ups. All follow-ups of the Raine Study were conducted in compliance with the Declaration of Helsinki and approved by the UWA HREC, and participants provided written informed consent for each follow-up prior to examination.

The Generation R study^10^ is a population-based prospective birth cohort study conducted in Rotterdam, The Netherlands. Women with a due data between April 2002 and January 2006, who were registered inhabitants in the municipality of Rotterdam at the time of delivery, and their offspring were eligible to participate in the Generation R study. The complete Generation R study has been approved by the Medical Ethical Committee of the Erasmus MC and conducted according to the World Medical Association Declaration of Helsinki. All examinations, interviews, and questionnaires were carried out after approval of the ethical committee, and after obtaining written informed consent from participants’ parent(s) or legal guardian(s).

The CLSA^11,12^ is a cohort study involving 51,338 individuals, aged 45–85 years at the time of enrolment. Baseline data collection was completed in 2015 and participants are re-examined every 3 years since. Study procedures were conducted in accordance with the World Medical Association Declaration of Helsinki ethical principles for medical research, and all participants provided informed written consent. The CLSA was approved by the University of Manitoba Health Research Ethics Board.

In Stage 2, we meta-analysed the Stage 1 cohorts with the UK Biobank previously reported by Zekavat et al.^14^ The Stage 2 meta-analysis summary statistics was used for post-GWAS analyses, PGS testing, and MR.

The PGS test cohorts comprised a European sample and an East Asian sample. The European cohort is a collective of 4 smaller studies: the Kidskin Young Adult Myopia Study,^63^ WA Eye Protection Study,^64^ WA Twin Eye Study, and a second smaller batch of the Raine Study Gen2 not included in the GWAS. The former three form the Ophthalmic WA Biobank (OWAB). The second batch of the Raine Study Gen2 cohort was genotyped and analysed separately from the rest of the sample due to a later whole blood sample collection. Eye examinations for all studies were conducted in 2009–2020 at the Lions Eye Institute, Nedlands, Western Australia. All participants 18+ years old at the time of participation provided written informed consent, while verbal assent was provided by participants aged <18 years along with written informed consent from their carers. All studies were conducted in accordance with the Declaration of Hensinki and approved by the UWA HREC.

The Beijing Eye Study is a population-based study conducted across 7 communities in Greater Beijing. All individuals living in these communities aged 40+ years were eligible. The baseline survey was conducted in 2001, with follow-ups conducted in 2006 and 2011.

OCT imaging of the choroid was conducted at the 2011 follow-up. All study follow-ups were approved by the Beijing Tongren Hospital Medical Ethics Committee, and all participants gave written informed consent for participation.

Cohort-specific details for the GWAS and PGS testing, including phenotyping and genomic data quality control are described below, while demographic information are described in **Supplementary Table 1 cohorts**.

### Phenotyping

All cohorts, except the Rotterdam Study, GenR, CLSA, and the UK Biobank, used Spectral-domain OCT imaging (Heidelberg Engineering, Heidelberg Germany) with enhanced depth imaging mode^65^ to penetrate the chorioscleral interface. One horizontal and one vertical 30-degree (∼8.7 mm) B-scan centred on the fovea were taken with each averaged over 100 frames to improve the signal-to-noise ratio. Scans were exported and images analysed offline. For the AugUR, Gutenberg, and Beijing Eye studies, subfoveal ChTs were measured manually by experienced observers. For the Australian studies (the Raine Study, BHAS, and Ophthalmic Western Australia Biobank) scans were exported, segmented, and measured offline using a semi-automated custom image analysis program,^66^ and then corrected for magnification effects resulting from varying axial lengths. Programs for segmentation, measurement, and magnification correction were written on MATLAB which measured the ChT at every 10µm intervals within a 0.5mm-radius circle centred on the fovea. The mean ChT within the central 0.5mm foveal region was used as the main outcome in these studies.^5,52^

The Rotterdam and Generation R studies conducted horizontal and vertical 30-degree B-scans centred on the fovea using a Topcon 3D 1000 mk2 (Topcon 3D 2000, Topcon DRI Atlantis/Triton). Segmentation for the Rotterdam Study was done using an in-house deep learning model, based on nnUNetv2 and trained using manually annotated data of 180 participants (708 OCT b-scans), the average ChT of the 0.5mm central foveal region was calculated. For Generation R, the raw segmentation data from central 0.5mm-radius calculated. Magnification effects due to varying axial lengths were corrected using the Littman method.

In the CLSA, ChT was estimated from fundus photographs using a machine-learning program. The program was trained on 4,324 fundal images and 0.5mm-diameter subfoveal ChT measurements from the BHAS. This was then validated and tested in 1,086 and 622 BHAS samples, respectively. The machine-learning model yielded a R^2^ = 0.52, mean absolute error= 43.2µm, and root mean square error= 55.1µm, comparable to the performances of previously-reported deep-learning models for ChT.^67,68^

Eyes with pathology that significantly affects the macula were removed from analyses. These include, but not limited to, current or previous macular degeneration (myopic or age-related), polypoidal choroidal vasculopathy, moderate/severe diabetic retinopathy, choroidal neovascularisation, macular oedema, posterior uveitis, central serous chorioretinopathy, and genetic choroidal diseases (e.g., choroideremia, gyrate atrophy). These were excluded based on participant-reported information or if revealed during the eye examination. Only the UK Biobank GWAS did not exclude eyes with pathology. However, the authors noted that the prevalence of ocular disease is low; ∼1.1% of participants have AMD,^14^ which is likely the most common macular condition that could affect ChT.

If ChT was available for both eyes, the average value of both eyes was calculated and analysed; if not, the value for the remaining eye was used for analysis. To account for heterogeneity between cohorts, a rank-based inverse normal transformation was applied to the subfoveal ChT for the GWAS within each cohort.

### Quality control and imputation of genetic data

Further details on genotyping and imputation for each cohort are available in **Supplementary Table 1**. Prior to imputation, SNPs with call rate <97%, minor allele frequency < 0.01, or deviations from the Hardy-Weinberg equilibrium at p< 10e^-6^ were removed from the analysis. Individuals with ≥3% SNP missingness or relatedness pi-hat >0.2 were excluded. Ancestral outliers based on principal components 1 and 2 that deviate >6 standard deviations from the respective cohorts’ main ancestry (European or East Asian) were also excluded. Following imputation, SNPs with imputation quality score R^2^ <0.3 were further removed prior to each GWAS. This threshold was then increased to 0.6 prior to the meta-analysis.

### GWAS and meta-analysis

Separate linear regression GWAS was conducted for each cohort and was adjusted for population stratification (PCs 1–10), age, sex, and axial length or, if axial length was not available, spherical equivalent refractive error as a proxy for axial length. The latter was corrected for due to the heavy influence of axial length on ChT. For studies without axial length data, eyes that have undergone laser refractive surgery or cataract removal were further excluded as these significantly affect refractive error. However, this could not be guaranteed in the UK Biobank. The CLSA did not have refractive error or axial length data, and the GWAS summary statistics was therefore adjusted for refractive error post-hoc using multi-trait-based conditional and joint analysis using GWAS summary data (mtCOJO, v1.91.7 beta1).^69^ The refractive error GWAS summary statistics used for the adjustment was taken from Hysi et al.^13^

Prior to meta-analysis, quality-control of GWAS summary statistics was conducted using the steps and GWASinspector scripts as described by Ani et al.^70^ Meta-analysis using an inverse-variance weighted method with fixed effects was then conducted using METAL.^71^ SNPs available in <5% of the combined study sample were excluded from further analysis in the GWAS summary statistics. In the Stage 2 GWAS, across all genome-wide significant variants, I^2^>50% was only observed for 17 non-lead SNPs within one locus on chromosome 8 (representing only 10% of SNPs on that locus).

### Post-GWAS analyses

The summary statistics from the Stage 2 GWAS was fed into the FUMA platform v1.8.2 (Functional Mapping and Annotation of Genome-Wide Association Studies; https://fuma.ctglab.nl).^72^ Functional annotation and gene mapping of independently significant SNPs were conducted using the SNP2GENE module in FUMA, which also performed 3D chromatin interaction mapping and calculated CADD scores. Variants with CADD scores >12.37 are considered potentially pathogenic.^73,74^ GWAS variants were further mapped to genes via eQTL in the retina (EyeGEx data)^75^ 48 tissues within the GTEx v8 database used for the other tissues.^76^ Gene-based and gene-set (pathway) analyses were performed with MAGMA (Multi-marker Analysis of GenoMic Annotation) v1.08^77^ within the FUMA pipeline. Gene sets within MAGMA were curated from 9 sources (https://www.gsea-msigdb.org/gsea/msigdb/collection_details.jsp#C2), including BioCarta, KEGG Medicus, Pathway Interactions Database, Reactome, SigmaAldrich, Signaling Gateway, SuperArray SABiosciences, WikiPathways, and KEGG Legacy Sets.

### Polygenic risk score

Summary statistics from the Stage 2 GWAS underwent clumping using Plink 2.0 with parameters ‘–clump-r2 0.1‘ and ‘–clump-kb 250‘. Ten 10 PGS for ChT were based on independent SNPs at varying thresholds for the p-values as modelled by the GWAS, ranging from p<5e^-8^ (genome-wide significant level) to p<1 (all independent SNPs). When examined in the European test sample (**Supplementary Table 1 cohorts**), the PGS comprising only genome-wide significant SNPs had the strongest association with ChT and therefore used for analysis. The average ChT of both eyes for each participant was tested against the PGS in a linear regression model, adjusted for age, sex, genotyping batch, axial length, and the first 5 principal components. Variance explained by the PGS alone (incremental R^2^) was calculated using the package ‘boot‘ on R statistical software. The same analysis method was applied for the cross-trait analysis with axial length.

### Heritability and genetic correlation

SNP-based heritability and genetic correlation were estimated from Stage 2 GWAS summary statistics using LD Score Regression (LDSC) v1.01.0. Narrow-sense heritability and genetic correlation with refractive error and axial length were further estimated using the GCTA (genome-wide complex trait analysis v1.94.0) software^78^ and the ‘MCMCglmm‘ package on R. Using GCTA, a genomic relation matrix was constructed from the genetic data of the BHAS cohort, against which, ChT and axial length were analysed using GREML. This was repeated with ChT and SphE as the outcome measures. The ‘MCMCglmm‘ R package runs a Multivariate Generalised Linear Mixed Models (GLMM) on pedigree-based data. Models were adjusted for sex, age, and the first 10 principal components.

### Colocalisation

To test for shared causal variants between ChT and refractive error, AMD, or PACG, we performed a Bayesian colocalisation analysis using approximate Bayes factors,^79^ with the ‘coloc’ package on R. This method allows the use of p-values for colocalisation analysis where betas are not available, as in the case for Hysi et al.’s publicly available refractive error GWAS summary statistics.^13^ A harmonised dataset was created from the summary statistics from our and Hysi et al.’s GWAS by extracting SNPs present in both datasets and aligning effect/non-effect alleles. SNPs with effect allele frequencies differing by >10% between the 2 datasets and were removed. Genomic boundaries for each of the 33 lead SNPs were extracted from the FUMA output. Where a locus was very small (<50 SNPs) or large (>1Mb), variants within 500 kb of the lead SNPs were used to define the region instead. The harmonised dataset was subset by locus and fed into the ‘coloc.abf()‘ function. This computed a posterior probability that a given genomic region contains signals for both traits arising from the same causal variant (PP4). The same was done for colocalisation with AMD causal variants using the summary statistics from He et al.^39^

### Mendelian randomisation

We explored putative ChT causal relationship with refractive error, AMD, and PACG using two-sample Mendelian randomisation. As the latest GWAS meta-analyses of the 3 latter traits^13,20,39^ had substantial sample overlap with the current Stage 2 GWAS, summary statistics for refractive error was obtained from the Genetic Epidemiology Research on Adult Health and Aging (GERA) cohort (n= 59,094),^80^ those for AMD was obtained from the International AMD Genetics Consortium (IAMDGC; n= 16,144 cases and 17,832 controls; http://eaglep.case.edu/iamdgc_web/),^81^ and those for PACG from the Millian Veteran Program (n= 7,129 cases and 439,835 controls).^15^ As beta values were not provided in the AMD summary statistics, the beta coefficient and standard error for each variant was estimated using a method described and validated previously.^39,82^ First, z-scores for each variant in the summary statistics were calculated from the p-values and direction of association. We assume that the standard error of the beta is proportionate to a constant, calculated as:

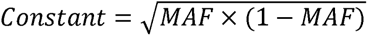

Where MAF= minor allele frequency. The MAF of each SNP in European populations was obtained from the 1000 genomes project. This constant was estimated using the average beta values from 52 variants from the IAMDGC publication,^81^ the odds ratios for AMD of which are available in GWAS Catalog (https://www.ebi.ac.uk/gwas/studies/GCST003219). This constant was then used to estimate beta coefficients and standard errors for the other SNPs.

We used Rstudio (version 4.4.2; The R Foundation for Statistical Computing Platform) and packages ‘TwoSampleMR’ and ‘MRPRESSO’ to conduct the analyses. For each exposure trait (ChT, refractive error, AMD, PACG), we filtered for genome-wide significant SNPs. These then underwent clumping to remove highly correlated variants, at a 0.1 r^2^ threshold and 10000kb genomic window. The common SNPs between the outcome and exposure datasets were then filtered and the datasets harmonised. Exposure–outcome causal relations were then analysed using 5 Mendelian randomisation methods – inverse variance weighted (IVW), MR-Eggar, simple mode, weighted mode, and weighted median – as the default within the TwoSampleMR package. The primary outcome was derived from the IVW analysis, while MR-Egger regression assessing for pleiotropy. For further sensitivity analysis, leave-one-out analysis and MR-PRESSO were performed.

## Data availability

The full summary statistics of the Stage 2 ChT GWAS meta-analysis can be obtained from the investigators who can be contacted at. The Hysi et al. refractive error GWAS summary statistics used in the colocalisation and LDSC analyses was downloaded from GWAS Catalog (study accession: GCST010003). The He et al. (2024) AMD summary statistics used in the colocalisation and LDSC analyses was obtained directly from the investigators who can be contacted at. The IAMDGC’s AMD summary statistics used in the MR analyses were downloaded from http://eaglep.case.edu/iamdgc_web/. The GERA’s refractive error GWAS summary statistics used in the MR analysis were obtained directly from the KP investigators who can be contacted at. The Verma et al. PACG GWAS summary statistics used in the colocalisation, LDSC, and MR analyses was downloaded from GWAS Catalog (accession code: GCST90475876). Data are available from the Canadian Longitudinal Study on Aging (www.clsa-elcv.ca) for researchers who meet the criteria for access to de-identified CLSA data.

## Funding

The Busselton Healthy Ageing Study (BHAS) is supported by grants from the Government of Western Australia (Department of Jobs, Tourism, Science, and Innovation), Western Australian Future Health Research and Innovation Fund (Grant IDs WACS-OSP:2023-2024, WACS-OSP:2025-2026), the City of Busselton, and private donations to the Busselton Population Medical Research Institute. General infrastructure support for the BHAS is provided by the Western Australian Country Health Service.

The eye data collection for the Raine Study Gen2 20- and 28-year follow-ups were funded by the NHMRC (grants 1021105, 1126494, and 1121979), Australian Vision Research (formerly the Ophthalmic Research Institute of Australia), Alcon Research Institute, Lions Eye Institute, BrightFocus Foundation, Australian Foundation for the Prevention of Blindness, Canadian Institutes of Health Research, and the Heart Foundation (grant 102170). GWAS data collected at the Raine Study Gen2 14- and 17-year follow-ups was funded by the NHMRC (grants 572613, 403981, 1059711) and the Canadian Institutes of Health Research (MOP-82893). The core management of the Raine Study is funded by The University of Western Australia, Curtin University, The Kids Research Institute Australia, Women and Infants Research Foundation, Edith Cowan University, Murdoch University, The University of Notre Dame Australia and the Western Australian Future Health Research and Innovation Fund (Grant IDs WACSOSP2023-2024, WACSOSP2025/7).

The Rotterdam Study and Generation R study are is made possible by financial support from Erasmus Medical Center; Erasmus University; the Netherlands Organization for the Health Research and Development (ZonMw); the Research Institute for Diseases in the Elderly (RIDE); the Ministry of Education, Culture and Science, and the Ministry for Health, Welfare and Sports of the Netherlands; the European Commission (DG XII); and the Municipality of Rotterdam. Additional funding was provided by Oogfonds; Stichting voor Ooglijders; Stichting voor Blindenhulp; Rotterdamse Stichting Blindenbelangen; Henkes Stichting; Algemene Nederlandse Vereniging ter Voorkoming van Blindheid; and Landelijke Stichting voor Blinden en Slechtzienden.

The AugUR study analyses were supported by grants from the German Federal Ministry of Education and Research (BMBF 01ER1206, BMBF 01ER1507 to I.M.H.), by the Deutsche Forschungsgemeinschaft (DFG, German Research Foundation; HE 3690/7-1 and HE 3690/5-1 to I.M.H., BR 6028/2-1 to C.B.), by the National Institutes of Health (NIH R01 EY RES 511967 and RES516564 to I.M.H.), and by institutional budget (University of Regensburg). The funding sources were not involved in study design, data collection, analysis and interpretation of data, or preparation of the manuscript.

The Gutenberg Health Study is funded through the government of Rhineland-Palatinate (“tiftung Rheinland-Pfalz für Innovation”, contract AZ 961-386261/733), the research programs “Wissen schafft Zukunft” and “Center for Translational Vascular Biology (CTVB)” of the Johannes Gutenberg-University of Mainz, and its contract with Boehringer Ingelheim, TRON and PHILIPS Medical Systems, including an unrestricted grant for the Gutenberg Health Study.

Funding for CLSA is provided by the Government of Canada through the Canadian Institutes of Health Research (CIHR) under grant reference: LSA 94473 and the Canada Foundation for Innovation, as well as the following provinces, Newfoundland, Nova Scotia, Quebec, Ontario, Manitoba, Alberta, and British Columbia.

Genotyping of the GERA cohort was funded by a grant from the National Institute on Aging, National Institute of Mental Health, and National Institute of Health Common Fund (RC2AG036607). Support for GERA participant enrollment, survey completion, and biospecimen collection for the Research Program on Genes, Environment and Health was provided by the Robert Wood Johnson Foundation, the Wayne and Gladys Valley Foundation, the Ellison Medical Foundation, and Kaiser Permanente Community Benefit Programs.

This work was further supported by a Perth Eye Foundation Grant awarded by the Australian Vision Research, and a Lions Eye Institute Research Implementation Group Strategic Grant. SSYL is supported by a Western Australia Future Health Research and Innovation Emerging Leaders Fellowship.

DJVH is supported by the European Research Council (ERC) under the European Union’s Horizon Europe research and innovation programme (ERC Advanced Grant to C.C.W. Klaver, grant agreement No. 101098324).

APK is supported by a UK Research and Innovation Future Leaders Fellowship (MR/Y033930/1), an Alcon Research Institute Young Investigator Award and a Lister Institute for Preventive Medicine Award.

HC was supported by the National Eye Institute (NEI) (R01EY027004 and R01EY033010). SM and PG are each supported by a NHMRC Investigator Grant.

DAM is supported by a Stan Perron Charitable Foundation People’s Grant.

## Data Availability

All data produced in the present study are available upon reasonable request to the authors.

## Acknowledgements

The authors would like to thank the Busselton Healthy Aging Study participants, the Raine Study participants, and their families for their ongoing participation in the study. We also thank the Raine Study team for study coordination and data collection, as well as the National Health & Medical Research Council (NHMRC, Australia) and the Raine Medical Research Foundation for their long-term contribution to funding the Raine Study over the last 30 years.

The Generation R Study Programme is conducted by the Erasmus MC, University Medical Center Rotterdam in close collaboration with the Erasmus University Rotterdam and the city of Rotterdam. We gratefully acknowledge the contribution of all participants. The general design of Generation R Study is made possible by long-term financial support from Erasmus MC, University Medical Center Rotterdam, the Netherlands Organization for Health Research and Development (ZonMw) and the Dutch Ministry of Health, Welfare and Sport. We are grateful to the Kaiser Permanente Northern California members who have generously agreed to participate in the Kaiser Permanente Research Program on Genes, Environment, and Health.

This research was made possible using the data and biospecimens collected by CLSA. This research has been conducted using the CLSA dataset (Baseline Comprehensive Dataset version 4.0), under Application Number 190225. The CLSA is led by Drs. Parminder Raina, Christina Wolfson and Susan Kirkland. The authors gratefully acknowledge the time and commitment of the CLSA participants, without whom this research would not be possible. The opinions expressed in this manuscript are the author’s own and do not reflect the views of the Canadian Longitudinal Study on Aging.

The Pawsey Supercomputing Centre provided computation resources to carry out analyses required with funding from the Australian Government and the Government of Western Australia.

## Conflicts of interest

APK has acted as a paid consultant or lecturer to Abbvie, Aerie, Google Health, Heidelberg Engineering, Glaucore, Novartis, Qlaris Bio, Regeneron, Reichert, Santen, Seonix Bio, Tavo Biotherapeutics, Thea and Topcon. GL is employed by Ocumetra. SM is co-founder of, holds stock in and consults for Seonix Bio.

## Notes

### Author Declarations

The University of Western Australia Human Research Ethics Committee provided ethical approval for the Raine Study, Busselton Healthy Aging Study, and Ophthalmic Western Australia Biobank. The Ethics Committee of the University of Regensburg gave ethical approval for the AugUR Study. The Medical Ethics Committee of the Erasmus MC provided ethical approval for the Generation R and Rotterdam Studies. The Medical Ethics Commission of Rhineland-Palatinate provided ethical approval for the Gutenberg Health Study. The University of Manitoba Health Research Ethics Board provided ethical approval for the Canadian Longitudinal Study on Aging. The Beijing Tongren Hospital Medical Ethics Committee provided ethical approval for the Beijing Eye Study.

### Summary of Updates

Authors' names and minor spelling errors in acknowledgements section.

