## Supplementary material for "Genome-wide discovery reveals 30 loci for choroidal thickness and uncovers potential causal links with angle-closure glaucoma": Table

### ***Table 1. Replication of previous variants***

| **Study** | **Lead SNP** | **Chr** | **Base position^** | **Mapped gene(s)** | **Replication**  **(p-value)** |
| --- | --- | --- | --- | --- | --- |
| Zekavat et al.^15^ | rs10691759 | 1 | 61332906 | *NFIA* | Yes; proxy SNP  (p=1.7e-5)** |
|  | rs7565795 | 2 | 298269 | *SH3YL1; ACP1; FAM150B* | Yes (p= 2.6e-7)** |
|  | rs13094358 | 3 | 25411118 | *RARB* | Nominally (p= 0.008)* |
|  | rs13118211 | 4 | 82400811 | *RASGEF1B* | Yes; proxy SNP  (p= 0.0023)** |
|  | rs11382941 | 4 | 146096470 | *HHIP; ANAPC10; ABCE1; OTUD4* | No (p= 0.36) |
|  | rs34904738 | 5 | 158029804 | *-* | Yes  (p= 0.0021)** |
|  | rs3063819 | 6 | 22072350 | *-* | No (p= 0.059) |
|  | rs74661234 | 8 | 8558309 | *CLDN23* | No^#^ |
|  | rs61198364 | 8 | 9192297 | *-* | Yes; proxy SNP (p= 0.0022)** |
|  | rs281730 | 8 | 108617455 | *-* | No (p= 0.22) |
|  | rs56161499 | 8 | 108752864 | *-* | No (p= 0.47) |
|  | rs707739 | 9 | 79118903 | *GCNT1* | No (p= 0.90) |
|  | ss1388024686 | 12 | 6954864 | *GNB3; CDCA3; USP5; SPSB2; LRRC23* | Nominally (p= 0.049)* |
|  | rs17174862 | 14 | 70398728 | *-* | Yes (p= 4.4e-6) |
|  | rs8012912 | 14 | 70474207 | *SMOC1* | Nominally (p= 0.005)* |
|  | rs12913832 | 15 | 28365618 | *OCA2; HERC2* | Yes (p= 3.7e-16) |
|  | rs683922 | 15 | 35008676 | *-* | Yes (p= 8.1e-9) |
|  | rs35988217 | 15 | 79370521 | *RASGRF1* | Yes; proxy SNP (p=8.4e-5) |
|  | rs135173 | 22 | 39646201 | *PDGFB; AL031590.1* | Yes (p= 0.0021)** |
| Hosada et al.^8^ | rs3753394 | 1 | 196620917 | CFH | No (p= 0.31) |
|  | rs7792658 | 7 | 49602718 | VIPR2 | No (p= 0.49) |

### **Nominally replicated at p< 0.05****;*** ***Replicated at Bonferroni corrected level of p< 0.0026 (0.05 divided by 19)****;*** *^#^ Not replicated as SNP or proxy SNPs not found in Stage 1 GWAS****;*** *^Position in build 37*

***Table 2: Lead SNPs from Stage 2 meta-analysis (IGGC + GenR + CLSA + UK Biobank)***

| **Locus** | **Lead SNP** | **Chr** | **Base position*** | **Effect allele** | **Non-effect allele** | **Beta (RINT)** | **P** | **Functional annotation** | ***Closest gene*** | **CADD** | **Novel** | **Association with related traits^#^** |
| --- | --- | --- | --- | --- | --- | --- | --- | --- | --- | --- | --- | --- |
| 1 | rs12739000 | 1 | 61322058 | T | G | 0.038 | 1.53E-13 | intergenic | *NFIA* | 5.39 | No | Refractive error |
|  | rs1777548 | 1 | 61481337 | T | C | -0.027 | 4.52E-08 | intronic |  | 1.38 |  |  |
| 2 | rs1329427 | 1 | 196704559 | T | C | 0.034 | 4.09E-12 | intronic | *CFH* | 0.67 | Yes | AMD |
| 3 | rs300751 | 2 | 209063 | A | C | 0.046 | 5.72E-21 | intergenic | *AC079779.7* | 0.66 | Yes | Refractive error |
| 4 | rs851382 | 2 | 19457282 | C | G | -0.122 | 5.06E-09 | ncRNA_intronic | *AC092594.1* | 1.32 | Yes |  |
| 5 | rs7625348 | 3 | 46044173 | T | C | -0.034 | 1.94E-09 | intergenic | *FYCO1* | 2.89 | Yes |  |
| 6 | rs144184445 | 3 | 187020711 | A | T | 0.155 | 3.95E-08 | intergenic | *MASP1* | 4.76 | Yes |  |
| 7 | rs13118211 | 4 | 82400811 | A | G | -0.029 | 1.15E-08 | intronic | *RASGEF1B* | 1.59 | No | Refractive error |
| 8 | rs11734405 | 4 | 146081096 | T | C | -0.027 | 2.89E-08 | intronic | *OTUD4* | 3.12 | No |  |
| 9 | rs2434612 | 5 | 158022041 | A | G | 0.054 | 1.28E-19 | intergenic | *RP11-32D16.1* | 3.43 | No |  |
| 10 | rs1383017982 | 6 | 22072350 | A | ACATC | 0.048 | 1.14E-12 | ncRNA_intronic | *CASC15* | 2.52 | Yes | Refractive error; corneal biometrics |
| 11 | rs6919851 | 6 | 151294962 | A | G | 0.041 | 7.82E-14 | intronic | *MTHFD1L* | 0.19 | Yes | Refractive error |
| 12 | rs11761083 | 7 | 156044166 | A | G | 0.030 | 1.38E-09 | intergenic | *RP11-362B23.1* | 2.26 | Yes |  |
| 13 | rs74661234 | 8 | 8558309 | T | C | 0.470 | 1.00E-08 | intergenic | *CLDN23* | 1.29 | Yes |  |
| 14 | rs13271489 | 8 | 9803712 | T | C | 0.030 | 1.41E-09 | intergenic | *snoU13 / XKR6* | 7.10 | Yes | Pathological myopia |
|  | rs11250098 | 8 | 10818607 | A | G | 0.028 | 1.93E-08 | intronic |  | 4.57 |  |  |
| 15 | rs199884711 | 8 | 108568628 | CT | C | -0.056 | 5.66E-13 | intergenic | *ANGPT1* | 1.06 | No |  |
| 16 | rs12285584 | 11 | 88935088 | T | C | -0.030 | 5.70E-09 | intronic | *TYR* | 3.20 | Yes | Inner retinal layers; intraocular pressure; hair/skin pigmentation; cataracts |
| 17 | rs571856 | 11 | 128691976 | T | C | -0.031 | 3.41E-08 | intergenic | *FLI1* | 5.27 | Yes | Refractive error |
| 18 | rs5442 | 12 | 6954864 | A | G | -0.065 | 2.64E-11 | exonic | *GNB3 / CDCA3* | **26.50** | No | Refractive error; retinal vasculature; inner retinal layers |
| 19 | rs770385 | 13 | 51142969 | A | G | 0.031 | 2.75E-10 | intergenic | *DLEU1* | 5.81 | Yes |  |
| 20 | rs7140683 | 14 | 70382672 | T | G | -0.029 | 4.15E-09 | intronic | *SMOC1* | 0.77 | No | - |
|  | rs17174862 | 14 | 70398728 | T | C | 0.063 | 7.01E-16 | intronic |  | 0.69 |  |  |
|  | rs12889195 | 14 | 70469131 | A | G | 0.036 | 1.22E-11 | intronic |  | 7.50 |  |  |
| 21 | rs10143492 | 14 | 75212114 | C | G | 0.029 | 3.60E-09 | intergenic | *FCF1* | 1.91 | Yes | - |
| 22 | rs12913832 | 15 | 28365618 | A | G | -0.075 | 9.08E-35 | intronic | *HERC2* | **15.81** | No | AMD; retinal vasculature; refractive error; corneal biometrics; hair/skin/eye colour |
| 23 | rs683922 | 15 | 35008676 | T | C | 0.040 | 6.37E-16 | intergenic | *GJD2* | 2.80 | No | Refractive error |
| 24 | rs6495367 | 15 | 79375347 | A | G | -0.037 | 2.72E-13 | intronic | *RASGRF1* | 3.24 | Yes | Refractive error |
| 25 | rs875391 | 15 | 89735582 | A | G | 0.032 | 1.02E-10 | intronic | *ABHD2* | 1.23 | Yes | Refractive error |
| 26 | rs2908972 | 17 | 11407259 | A | T | -0.028 | 2.41E-08 | intronic | *SHISA6* | **15.71** | Yes | Refractive error |
| 27 | rs34609292 | 17 | 12561079 | A | G | 0.035 | 1.25E-11 | intergenic | *MYOCD* | 0.49 | Yes |  |
| 28 | rs13381225 | 18 | 42812127 | A | G | -0.043 | 7.30E-12 | intronic | *SLC14A2* | 2.26 | Yes | Refractive error |
|  | rs1462151 | 18 | 42896817 | C | G | -0.040 | 7.71E-09 | intronic |  | 8.05 |  |  |
| 29 | rs12953562 | 18 | 47370934 | T | C | 0.043 | 6.18E-11 | ncRNA_intronic | *SCARNA17 / MYO5B* | 1.43 | Yes | Refractive error |
| 30 | rs135173 | 22 | 39646201 | T | C | -0.034 | 1.19E-09 | intergenic | *PDGFB* | **14.47** | No | AMD |

*AMD= age-related macular degeneration; CADD= Combined annotation dependent depletion score. SNPs with values >12.37 (bolded) are considered deleterious; RINT= rank-inverse transformed. *Position in build 37; ^#^ Selected associations for locus (not closest gene) that are potentially relevant to choroidal thickness*

**Table 3: Significant genes from MAGMA^**

| **Gene** | **Chr** | **Position start*** | **Position end*** | **#SNPs** | **Z-statistics** | **P** | **Annotated to GW-significant SNPs** |
| --- | --- | --- | --- | --- | --- | --- | --- |
| *LMX1A* | 1 | 165161104 | 165335952 | 940 | 4.86 | 5.75E-07 | No |
| *CFH* | 1 | 196611008 | 196726634 | 533 | 7.24 | 2.26E-13 | Yes |
| *CFHR2* | 1 | 196778898 | 196938356 | 806 | 5.11 | 1.59E-07 | No |
| *CFHR4* | 1 | 196809371 | 196898102 | 431 | 5.51 | 1.76E-08 | No |
| *SH3YL1* | 2 | 207730 | 276398 | 263 | 6.37 | 9.15E-11 | Yes |
| *ACP1* | 2 | 254140 | 288283 | 123 | 6.11 | 5.00E-10 | No |
| *FAM150B* | 2 | 269558 | 298851 | 107 | 6.11 | 5.00E-10 | No |
| *RARB* | 3 | 25205823 | 25649423 | 2914 | 4.60 | 2.15E-06 | No |
| *FYCO1* | 3 | 45949396 | 46047316 | 436 | 4.70 | 1.32E-06 | Yes |
| *HHIP* | 4 | 145557173 | 145676423 | 364 | 5.00 | 2.93E-07 | No |
| *ANAPC10* | 4 | 145878264 | 146029693 | 359 | 5.79 | 3.57E-09 | No |
| *OTUD4* | 4 | 146021990 | 146111313 | 220 | 4.72 | 1.18E-06 | Yes |
| *MTHFD1L* | 6 | 151176685 | 151433023 | 1699 | 6.59 | 2.25E-11 | No |
| *TNKS* | 8 | 9403424 | 9649856 | 1524 | 4.72 | 1.21E-06 | No |
| *MSRA* | 8 | 9901778 | 10296401 | 2805 | 6.33 | 1.21E-10 | No |
| *RP1L1* | 8 | 10453859 | 10579697 | 747 | 5.33 | 4.81E-08 | Yes |
| *XKR6* | 8 | 10743555 | 11068875 | 1964 | 5.78 | 3.84E-09 | No |
| *C8orf12* | 8 | 11215911 | 11306167 | 740 | 4.82 | 7.36E-07 | No |
| *BLK* | 8 | 11341510 | 11432113 | 777 | 4.92 | 4.33E-07 | No |
| *SMOC1* | 14 | 70310848 | 70509083 | 1128 | 7.90 | 1.42E-15 | Yes |
| *YLPM1* | 14 | 75220069 | 75332244 | 467 | 5.24 | 8.24E-08 | Yes |
| *DLST* | 14 | 75338594 | 75380448 | 218 | 4.70 | 1.33E-06 | No |
| *HERC2* | 15 | 28346186 | 28577298 | 630 | 6.28 | 1.68E-10 | No |
| *RASGRF1* | 15 | 79242289 | 79393115 | 713 | 6.55 | 2.97E-11 | Yes |
| *ABHD2* | 15 | 89620690 | 89755591 | 576 | 5.11 | 1.57E-07 | Yes |
| *RLBP1* | 15 | 89743100 | 89774982 | 165 | 6.11 | 5.00E-10 | Yes |
| *SLC14A2* | 18 | 42782960 | 43273072 | 2698 | 5.47 | 2.21E-08 | No |
| *BCL2* | 18 | 60780579 | 60997361 | 1035 | 4.93 | 4.16E-07 | Yes |
| *AL031590.1* | 22 | 39646865 | 39667063 | 132 | 4.69 | 1.35E-06 | No |

*MAGMA= Multi-marker Analysis of Genomic Annotation; GW= genome-wide.*

**Position in build 37; ^Significant at Bonferroni-corrected level of p< 2.48e-6, based on testing of 20175 mapped genes*
