## Supplementary Figure for "Genome-wide discovery reveals 30 loci for choroidal thickness and uncovers potential causal links with angle-closure glaucoma"

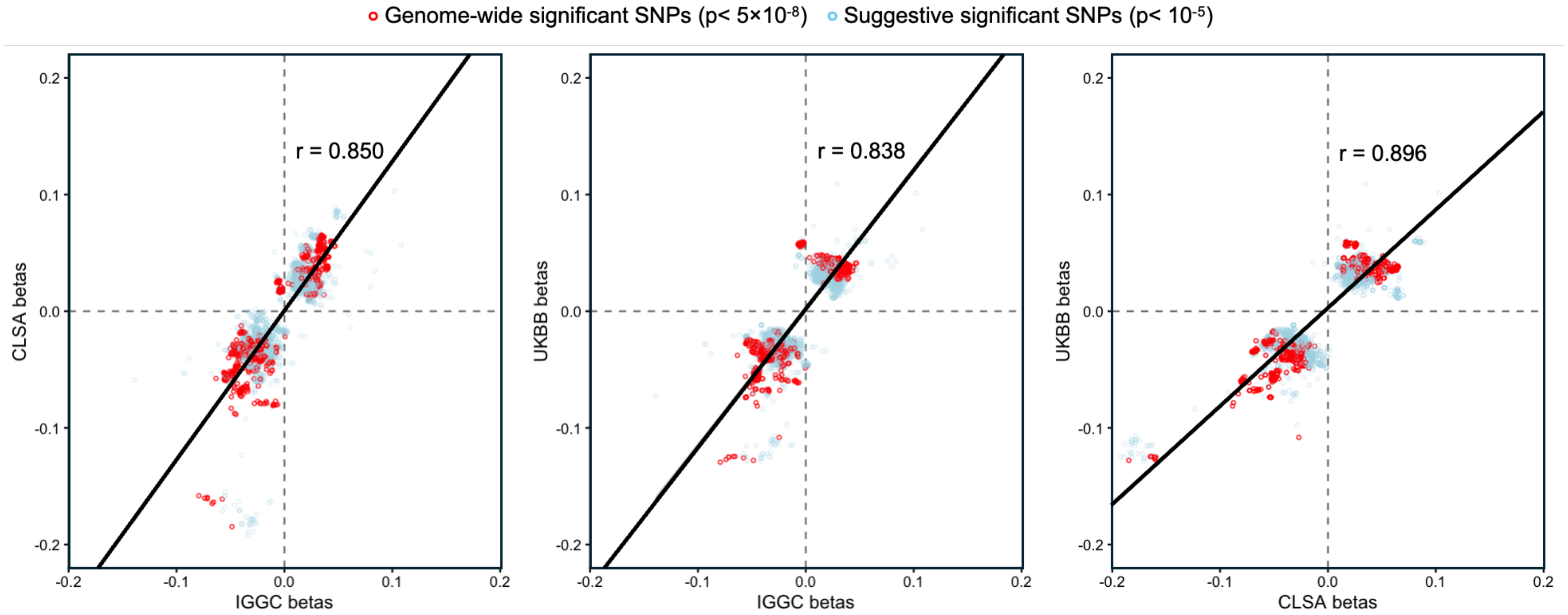


***Supplementary Figure 1.*** *Correlation in effect sizes of 4,690 SNPs that reach suggestive ($p< 10^{-5}$) or genome-wide ($p< 5\times10^{-8}$) significance between the IGGC+GenR (n= 15,416), CLSA (n= 18,443), and UK Biobank (n=44,823). Only ~3% of SNPs with genome-wide or suggestive significances had inconsistent direction of associations between the IGGC and the other 2 cohorts.*

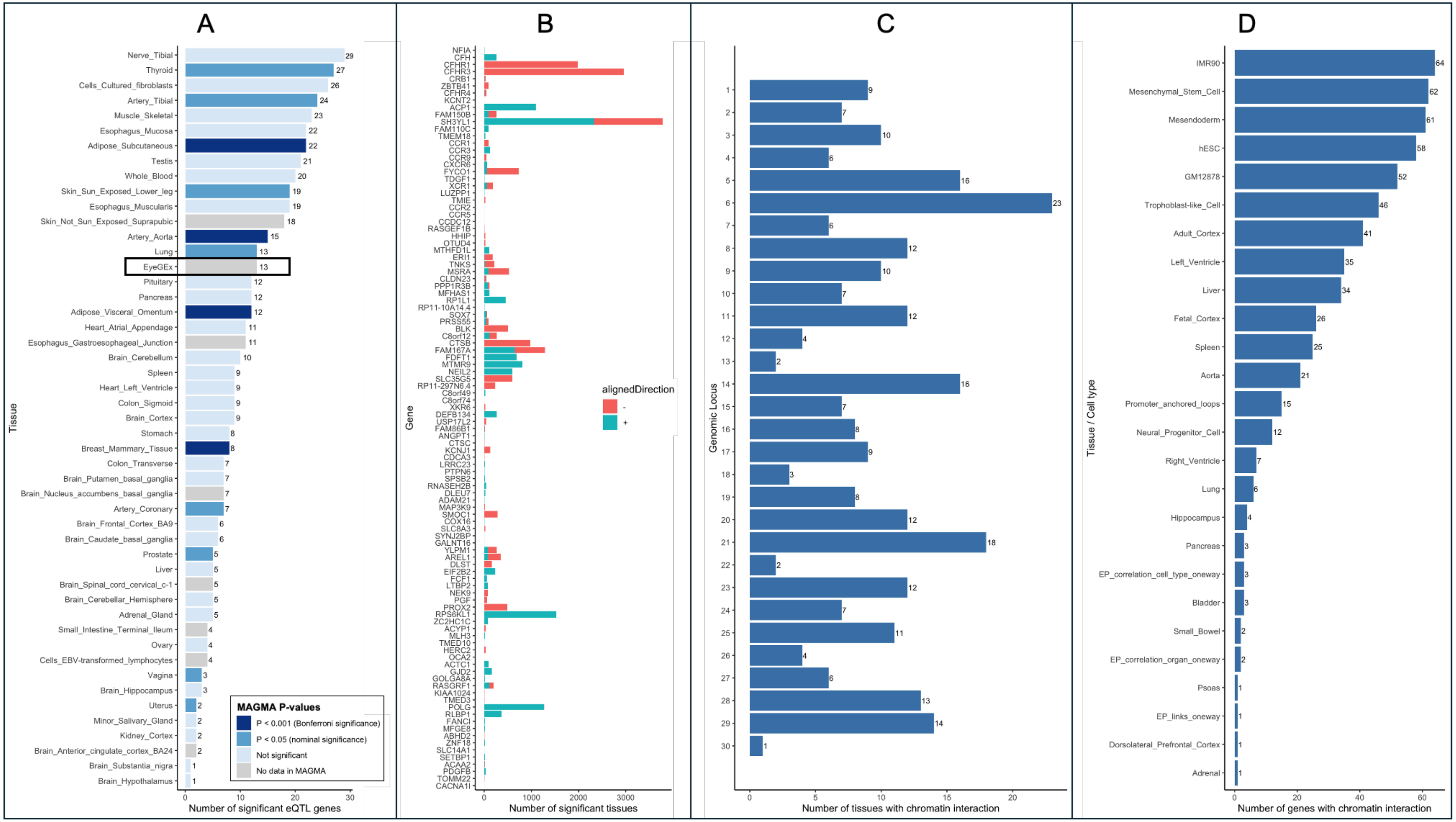


***Supplementary Figure 2****. Panel A: Number of significant eQTL genes per tissue, across the 48 tissues in GTEx version 8 plus the retina (EyeGEx; in black rectangle as the most relevant tissue), colour-coded based on MAGMA p-value for enrichment of GWAS signal for ChT within genes specifically expressed in each tissue. Retinal (EyeGEx) and some GTEx v8 tissues were not included in the MAGMA gene expression reference panel and therefore were not tested for enrichment. Panel B: Number of significant eQTL tissues (48 in GTEx v8 plus EyeGEx) per gene identified in the GWAS. Bars are colour coded based on direction of association between gene expression in each tissue and ChT. Size of blue bars represent number of genes where a higher expression in that tissue is linked with an increase in ChT, while size of red bars represents number of genes where a higher expression in that tissue is linked with a decrease in ChT. Panel C: Number of tissues with significant chromatin interaction per genomic locus. Panel D: Number of genes with significant chromatin interaction per tissue.*

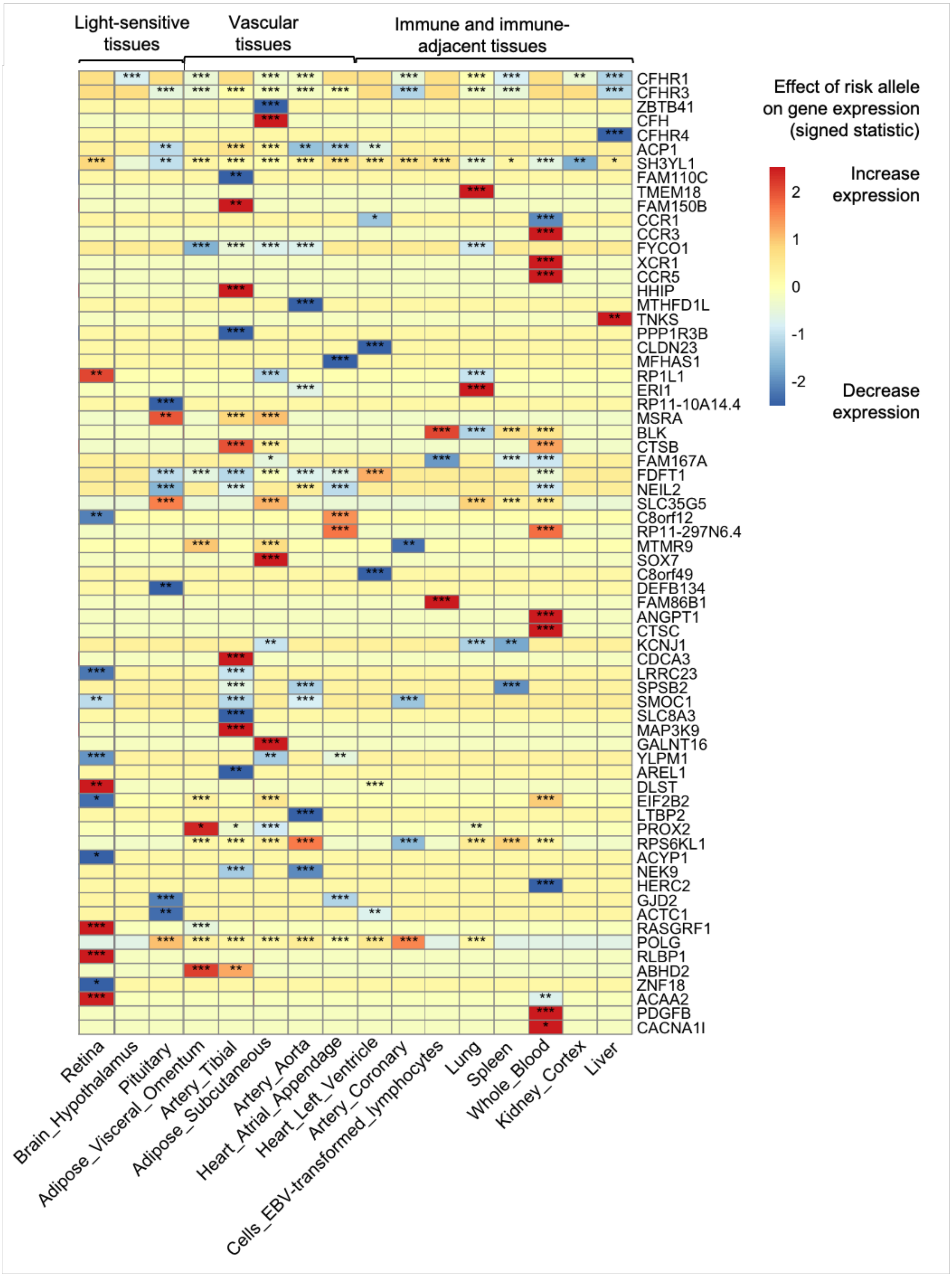


***Supplementary Figure 3***. *Heatmap of significant eQTL genes across 16 selected tissues of interest relevant to the ChT. Significant at *false discovery rate (FDR) < 0.05, **FDR< 0.01, or ***FDR< 0.001.*

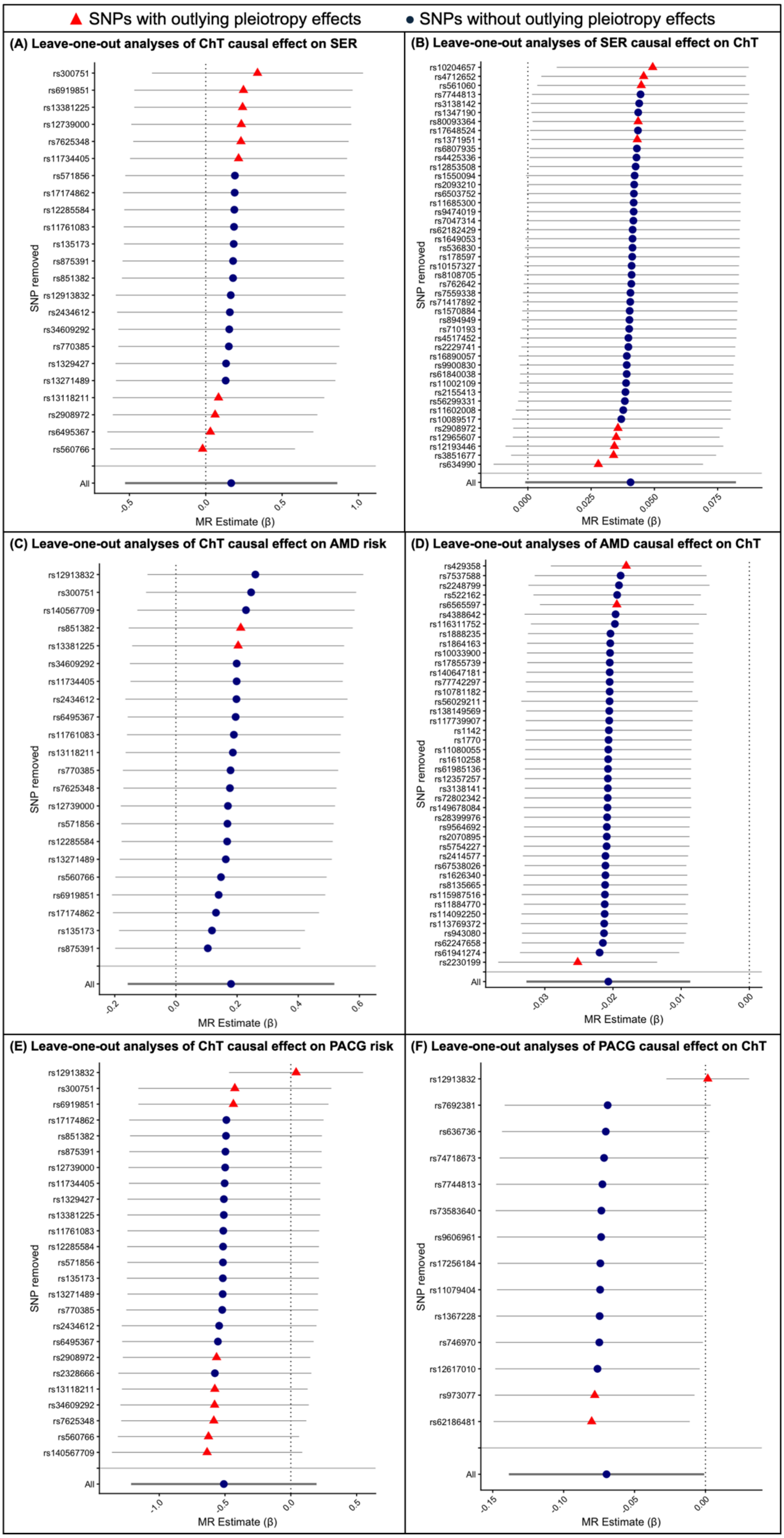


***Supplementary Figure 4.*** *Leave-one-out sensitivity analyses of putative causal effect of (A) ChT on refractive error, (B) refractive error on ChT, (C) ChT on AMD risk, (D) AMD on ChT, (E) ChT on PACG risk, and (F) PACG on ChT. Error bars show 95% confidence interval. Pleiotropy effects determined using MR-PRESSO. AMD= age-related macular degeneration; ChT= choroidal thickness; SER= spherical equivalent refractive error.*
